# Estimating clinical risk in gene regions from population sequencing cohort data

**DOI:** 10.1101/2023.01.06.23284281

**Authors:** James D. Fife, Christopher A. Cassa

**Affiliations:** Division of Genetics, Brigham and Women’s Hospital, Harvard Medical School, Boston, Massachusetts; Harvard Medical School, Boston, Massachusetts

## Abstract

While pathogenic variants significantly increase disease risk in many genes, it is still challenging to estimate the clinical impact of rare missense variants more generally. Even in genes such as *BRCA2* or *PALB2*, large cohort studies find no significant association between breast cancer and rare germline missense variants collectively. Here we introduce REGatta, a method to improve the estimation of clinical risk in gene segments. We define gene regions using the density of pathogenic diagnostic reports, and then calculate the relative risk in each of these regions using 109,581 exome sequences from women in the UK Biobank. We apply this method in seven established breast cancer genes, and identify regions in each gene with statistically significant differences in breast cancer incidence for rare missense carriers. Even in genes with no significant difference at the gene level, this approach significantly separates rare missense variant carriers at higher or lower risk (*BRCA2* regional model OR=1.46 [1.12, 1.79], p=0.0036 vs. *BRCA2* gene model OR=0.96 [0.85,1.07] p=0.4171). We find high concordance between these regional risk estimates and high-throughput functional assays of variant impact. We compare with existing methods and the use of protein domains (Pfam) as regions, and find REGatta better identifies individuals at elevated or reduced risk. These regions provide useful priors which can potentially be used to improve risk assessment and clinical management.

## Introduction

Large cohort studies have identified numerous genes in which carriers of germline variation have an increased predispositional risk of developing breast cancer.^1^ In particular, pathogenic coding variants in these genes are associated with significantly increased risk, and are routinely screened in diagnostic testing panels.^2^ However, translating this risk to carriers of rare missense Variants of Uncertain Significance (VUSs) continues to pose a challenge in the diagnostic setting.

Collectively, many individuals harbor rare VUSs in predispositional cancer genes, potentially increasing their risk of disease. However, on the basis of the collective frequency of these variants, some are unlikely to be highly penetrant or functionally impactful.^3,4^ Given their low frequencies, it is challenging to assess their clinical significance using epidemiological evidence at the variant level.

One approach to improve risk assessment has been to define groups of missense variants, or to identify genic regions which may confer higher or lower risk. These methods include modeling the depletion of variation using population sequencing data (selective constraint) using sets of exons,^5^ protein domains and regulatory sequences,^6^ three dimensional protein structures,^7^ sequence contact sets,^8^ specific disease phenotypes,^9^ and evolutionary conservation data.^10^ These methods have excellent resolution in genes or regions under strong selection, even in genes whose function is not well established, but can otherwise be challenged by the effects of drift, population structure, or sampling variance at low allele counts.

Other approaches have identified regions which are enriched for pathogenic variation and depleted of putatively neutral variation, including at protein interfaces,^11^ within protein structures,^12^ across genes and gene families,^13^ in pathways,^14^ and homologous regions.^15^ These methods make use of known biological structures and abundant clinical diagnostic data, but may also be challenged by the biases of prior diagnostic observations (whether from case ascertainment or assessment process) and consequently may not reflect the relative risk in population screening.

The interpretation of germline variants in clinically actionable disease genes is becoming commonplace in biobanks and large population health studies. The American College of Medical Genetics and Genomics recommends a set of 78 genes (ACMG SF v3.1) be reviewed, regardless of the indication for sequencing.^16^ Many of these genes are associated with predispositional cancer risk, and are commonly screened in the diagnostic setting. The ACMG/Association for Molecular Pathology (AMP) guidelines for sequence variant interpretation consider several related forms of evidence, including mutational ‘hot spots’ or well-studied functional domains without benign variation (PM1) and high rates of known pathogenic and low rates of benign missense variants (PP2). Here, we develop a framework to improve the estimation of risk within gene regions for rare missense VUSs derived from clinical diagnostic and population health data. This approach may be useful toward identifying protein segments which confer higher or lower risk, as well as providing an informative prior probability of pathogenicity for variation within a region.

## Material and Methods

### Defining genic regions with significant differences in germline cancer risk

To define genic regions with potentially distinct clinical risk of breast cancer, we identify segments which are enriched or depleted in pathogenic variant reports in ClinVar.^17^ We restrict our analyses to missense variants, removing all stop-gain, frameshift, and canonical splice site variants. We then use Jenks natural breaks optimization to partition the transcript based on the coding positions of pathogenic or likely pathogenic (P/LP) breast cancer reports.^18^ We break each transcript into 15 regions, or the maximum number of regions possible while maintaining sufficient numbers of missense carriers in each region to make risk estimates (**Supplementary Methods, Supplementary Table 1**).

We then make use of clinical data from the UK Biobank (UKB) to estimate the risk attributable to carrying a rare missense variant in each region.^19^ First, we establish a baseline risk for individuals who carry any rare missense variant in each gene. We perform a univariate Cox regression, comparing the risk for carriers of any rare missense variant to non-carriers.^20^ The resulting partial hazard for missense carriers in each gene,, is used in further comparisons. We next calculate the risk of carrying a rare missense variant in each predefined protein region. The resulting partial hazard for each region,, is compared to, to identify an elevated or reduced clinical risk in each region **(Figure 1)**. The risk ratio is thresholded, defining a relative risk of 1.15 or above as a higher risk region (HRR) or 0.85 or below as a lower risk region (LRR).

**Figure 1.**
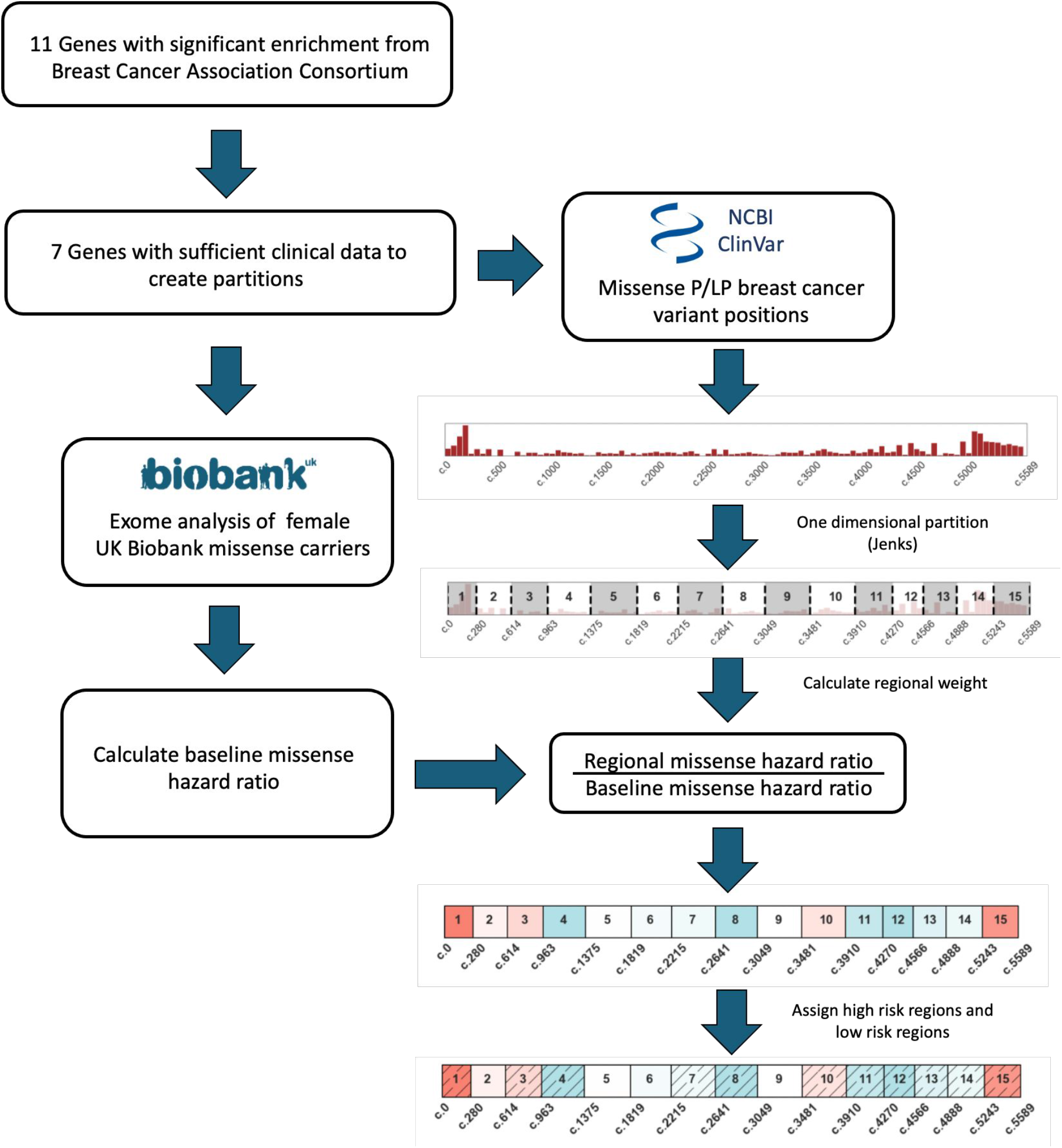
Assessing regional risk using clinical diagnostic and population sequencing data. We analyze data from 34 genes from a well-powered breast cancer meta-analysis (Breast Cancer Association Consortium) for their potential to have protein regions which confer higher or lower clinical risk for carriers of rare missense variants.^1^ We restrict to 7 genes with sufficient clinical diagnostic data in ClinVar, and use the distribution of pathogenic variant reports to partition each gene into distinct regions (Methods). Using those regional boundaries, we then use breast cancer status and population sequencing data from over 100,000 women in the UK Biobank to calculate the missense risk ratio in each region. Risk ratio values are thresholded to label sets of regions as high risk regions (HRR) or low risk regions (LRR), and we find that these ratios can significantly distinguish patients at elevated or reduced clinical risk. Such risk values may be aligned with clinical diagnostic guidelines (ACMG/AMP PM1, PP2), or added to integrative prediction methods.

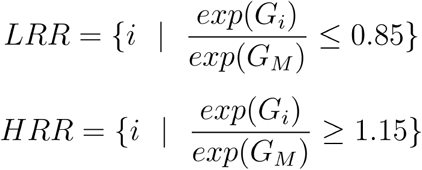

This threshold provides the highest effect size such that all seven genes evaluated have at least one high risk and one low risk region using this set of partitions **(Figure 2A, Supplementary Table 1-2)**.

**Figure 2.**
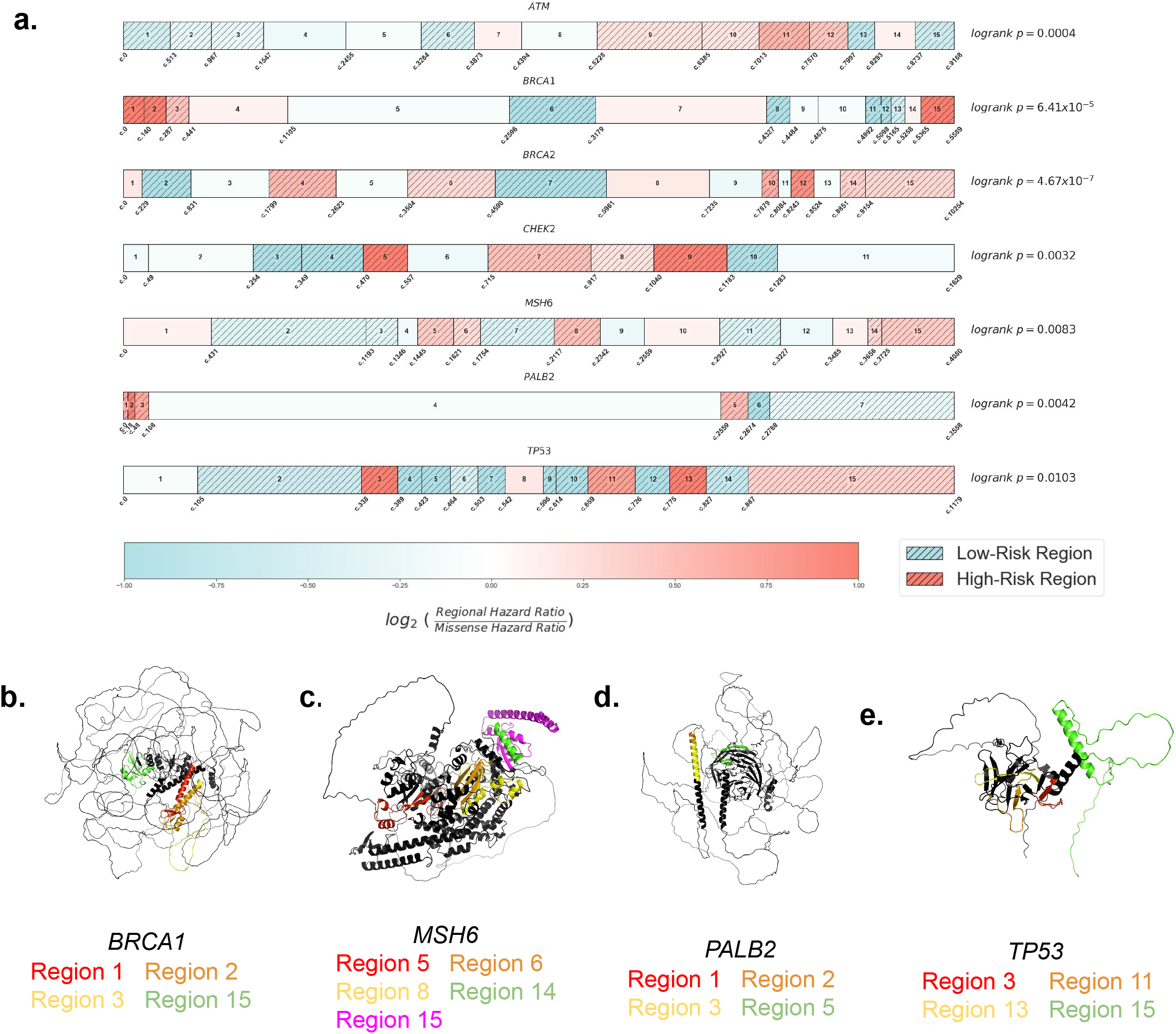
Regional partitions of breast cancer genes and structural distribution of high risk regions. =**[a]** Regional boundary definitions are calculated using the distribution of pathogenic variant reports in ClinVar and Jenks Natural Breaks optimization, spanning the entire length of each transcript (Methods). For each gene, high risk regions (HRR) are assigned for regions with risk ratios greater than or equal to 1.15, and low risk regions (LRR) for those less than or equal to 0.85. A relative risk ratio is computed as the Cox proportional hazard ratio of rare missense carriers in each region divided by the Cox proportional hazard ratio for rare missense carriers across all regions of each gene. Using breast cancer outcome data, we compare risk among rare missense carriers for variants in HRRs and LRRs (logrank p-values, right). **[b - e]** *BRCA1, MSH6, PALB2*, and *TP53* high risk regions highlighted on AlphaFold predicted protein structures. Despite the distance in one dimensional nucleotide sequence, HRRs are often aligned in three dimensional space.

### Exome sequencing and variant annotation

Exome sequencing was performed for UKB participants as previously described.^19^ Variant allele frequencies were estimated from the Genome Aggregation Database (gnomAD v2.1 exomes N=125,748). Variants were included with population maximum allele frequencies of ≤ 0.005 (Ensembl gnomAD plugin)^21^ or if not present in gnomAD. The canonical functional consequence of each variant was calculated using Variant Effect Predictor (v99) and we restrict our relative risk calculations to missense variants.^22^ For effect size comparison analyses, we specify predicted loss of function (pLOF) variants to include frameshift, stop gain, canonical splice-site, start lost, and stop lost annotation, and separately analyze synonymous variants. Non-coding variants outside of essential splice sites were not considered in the analysis. Variants which are non-PASS filter quality in gnomAD were excluded, as well as any variants in low complexity regions, segmental duplications, or other regions known to be challenging for next generation sequencing alignment or calling.^23^

### Selecting genes for analyses

Drawing from 34 putative cancer predisposition genes evaluated by the Breast Cancer Association Consortium,^1^ we retained genes with either statistically significant differences in breast cancer for carriers of rare non-synonymous variants, or genes with large effect sizes (odds ratio ≥ 2), resulting in 11 genes. We removed 4 of these genes (*BARD1, PTEN, RAD51D, RAD51C*) either due to a lack of P/LP missense reports in ClinVar, which prevented derivation of regional boundaries, or a lack of variant carriers with breast cancer in the UKB, which prevented reliable risk estimates in each region. For the remaining 7 genes in this study, we restrict to P/LP ClinVar reports annotated for breast or ovarian cancer with the exception of *MSH6* where we do not restrict to breast or ovarian cancer reports.

### Selecting parameters for regional boundary definitions

We selected a relative risk threshold for missense carriers of ± 0.15, as it was the largest tested value such that all seven genes had at least one high and low risk region. The optimization approach to define regional boundaries from pathogenic reports requires setting a number of regions *a priori*, which was initially set to 15 for all genes. When a gene could not be broken into 15 regions reliably and with sufficient power (carriers and cases) to perform Cox regressions without convergence errors, we selected the maximum number of regions which allowed such regressions to converge.

### Participant exclusion criteria

Males were excluded from all analyses, as well as individuals with missing information regarding patient age, or age of incident or prevalent cancer diagnosis. Analyses were performed separately for each gene.

Individuals with multiple missense variants in the same gene were excluded from analysis. Individuals with an LOF variant were grouped in the LOF category for each gene. For each gene analysis, we removed individuals who carry LOF variants in any of the other six genes if LOF variants in that gene are known to have a significant difference in breast cancer risk as measured by logrank p-value (p ≤ 0.05).

## Results

### Regional partitions identify gene segments which confer varying levels of risk

Using this approach, we define regional boundaries and calculate relative risks for carriers of rare missense variants, and identify regions which confer higher (HRR) and lower risk (LRR). We find that carriers of variants in HRRs have a significantly different risk of breast cancer from carriers of variants in LRRs in all 7 genes analyzed (**Figure 2A**). We find that regions of elevated risk cluster closely in three dimensional space in several genes, despite substantial distance in the one-dimensional transcript sequence. Using AlphaFold structural predictions available for five genes,^24^ we find multiple HRRs within *BRCA1* (Regions 1-3 vs. 15), *MSH6* (Region 8 vs. 14 and 15), *PALB2* (Regions 1-3 vs. 5), and *TP53* (Region 3 vs. 11 and 13) in close three dimensional proximity but far apart in the nucleotide sequence (**Figure 2B-D**). HRRs in *CHEK2* were adjacent in the protein core and were also adjacent in one dimensional space (**Supplementary Figure 1**).

### Regional information improves clinical risk prediction

Consistent with findings from a large meta-analysis examining breast cancer incidence,^1^ we find that rare missense variants do not always confer a significantly increased risk of breast cancer. Only 2 of the 7 genes confer an increased risk at the gene level for rare missense variant carriers: *ATM* (O.R. = 1.17 [1.06, 1.28], logrank p = 0.011) and *CHEK2* (O.R = 1.37 [1.19, 1.56], logrank p = 4.93×10^−5^), and other well-known genes such as *BRCA2* have no significant difference in risk (O.R = 0.95 [0.79, 1.12], logrank p = 0.33).

In contrast, REGatta can distinguish higher and lower risk for carriers of rare missense variation. We find significant differences in breast cancer incidence among missense carriers in HRRs vs LRRs in all seven genes analyzed (logrank p < 0.05) (**Supplementary Figure 2**). Observed effect sizes for HRR carriers vs. LRR carriers exceed those observed when comparing missense carriers vs. non-carriers in all seven genes (**Figure 3A, Supplementary Table 3**).

**Figure 3.**
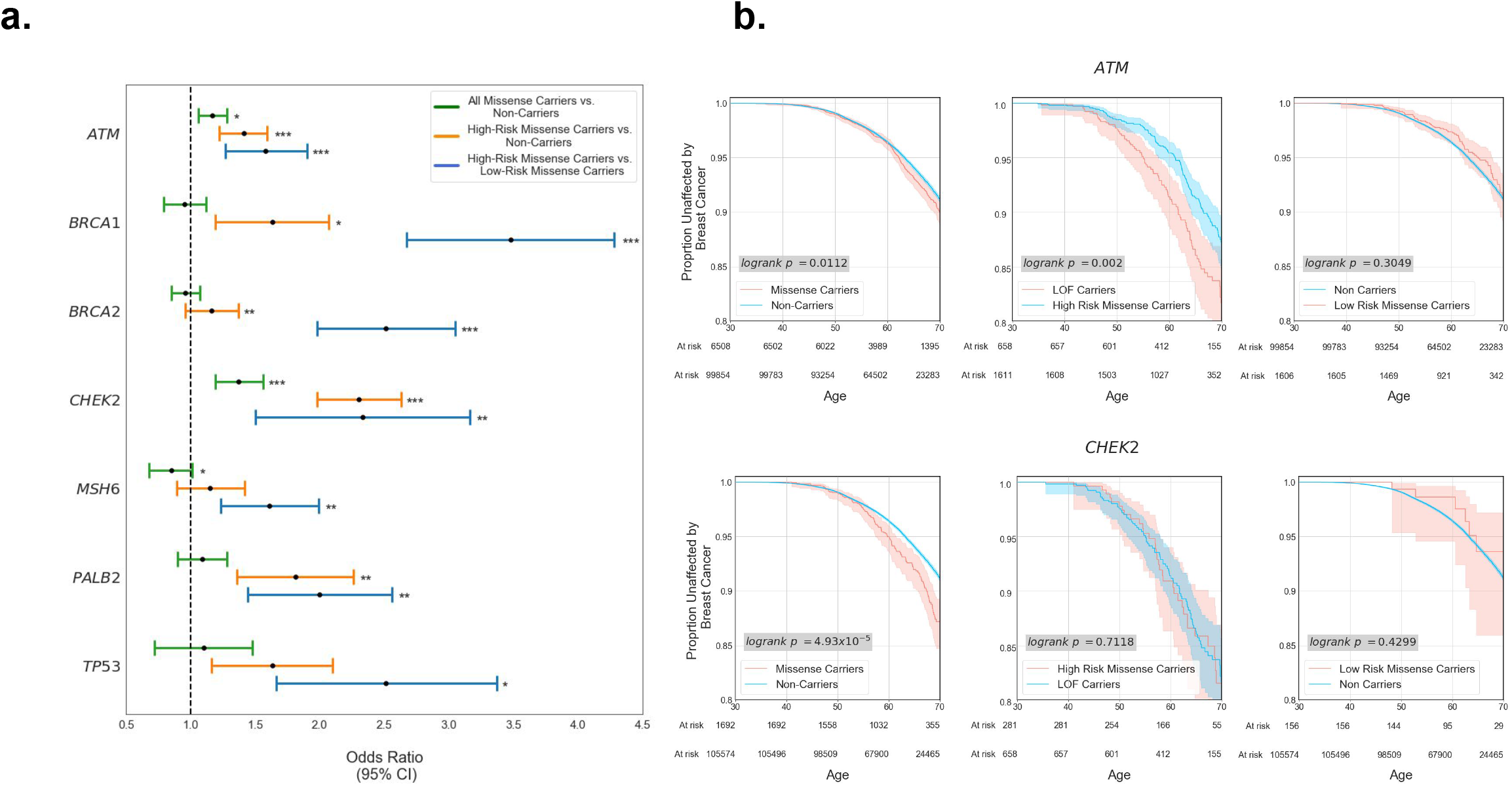
Regional stratification improves specificity in elevated risk predictions. **[a]** Age 65 Odds Ratio (OR) for the seven genes partitioned into high risk regions (HRR) and low risk regions (LRR) for rare missense variants. At the gene level, only two genes have an increased risk at the gene level for missense variant carriers when compared to non-carriers (green bars: *CHEK2, ATM*). Partitioning each gene transcript into HRR and LRR regions results in significant differences in breast cancer incidence among rare missense variant carriers in all 7 genes examined (blue bars). **[b]** Kaplan-Meier curves of patient breast cancer outcomes. While missense variants in aggregate in *ATM* and *CHEK2* are significantly associated with breast cancer at the gene level, patients who carry variants in LRRs have no significant difference in risk when compared with non-carriers in both genes. Additionally, carriers of rare missense variants in HRRs within *CHEK2* have no significant difference in breast cancer incidence from those who carry a predicted loss of function (canonical splice site, stop gain, or frameshift) variant.

This extends to genes which confer a significant risk at the gene level for rare missense carriers (*ATM, CHEK2*): REGatta distinguishes regions with higher and lower relative risk. Regions which are predicted as LRR have no significant difference in breast cancer incidence when compared with non-carriers in *ATM* (O.R. = 0.89 [0.63, 1.15], logrank p = 0.30) or *CHEK2* (O.R. = 0.99 [0.22, 1.75], logrank p =0.43). Importantly this distinguishes regions where rare missense variants are unlikely to substantially increase clinical risk in genes where they generally confer significantly increased risk (**Figure 3B**). Further, in *CHEK2*, we find no significant difference in breast cancer incidence among HRR missense carriers and predicted loss-of-function variant carriers (O.R. = 0.95 [0.56, 1.34], logrank p = 0.71) (**Figure 3B**), emphasizing the utility of this approach in identifying missense variation with large estimated functional impacts.

### Validation of regional risk assessments using functional assay data

Measurements from well-established functional assays may be considered strong evidence of pathogenicity in the clinical variant assessment process (PS3).^25^ We make use of experimental evidence from high-throughput variant installation assays as an orthogonal source of validation for our regional risk assessments.^26–31^ For the four genes where functional impact is reported on a continuous scale, we find significant differences in assay measurements between variants in HRRs vs LRRs (Kolmogorov–Smirnov p < 0.05, **Figure 4A**). In two genes where functional impact is reported dichotomously (either damaging or neutral), we find a statistically significant difference in *CHEK2* HRR vs. LRR reports (Enrichment = 2.61, ^2^ p = 0.003) and an increased but non-significant effect size in *MSH6* HRR vs. LRR reports (Enrichment = 1.24, ^2^ p = 0.06, **Figure 4B**).

**Figure 4.**
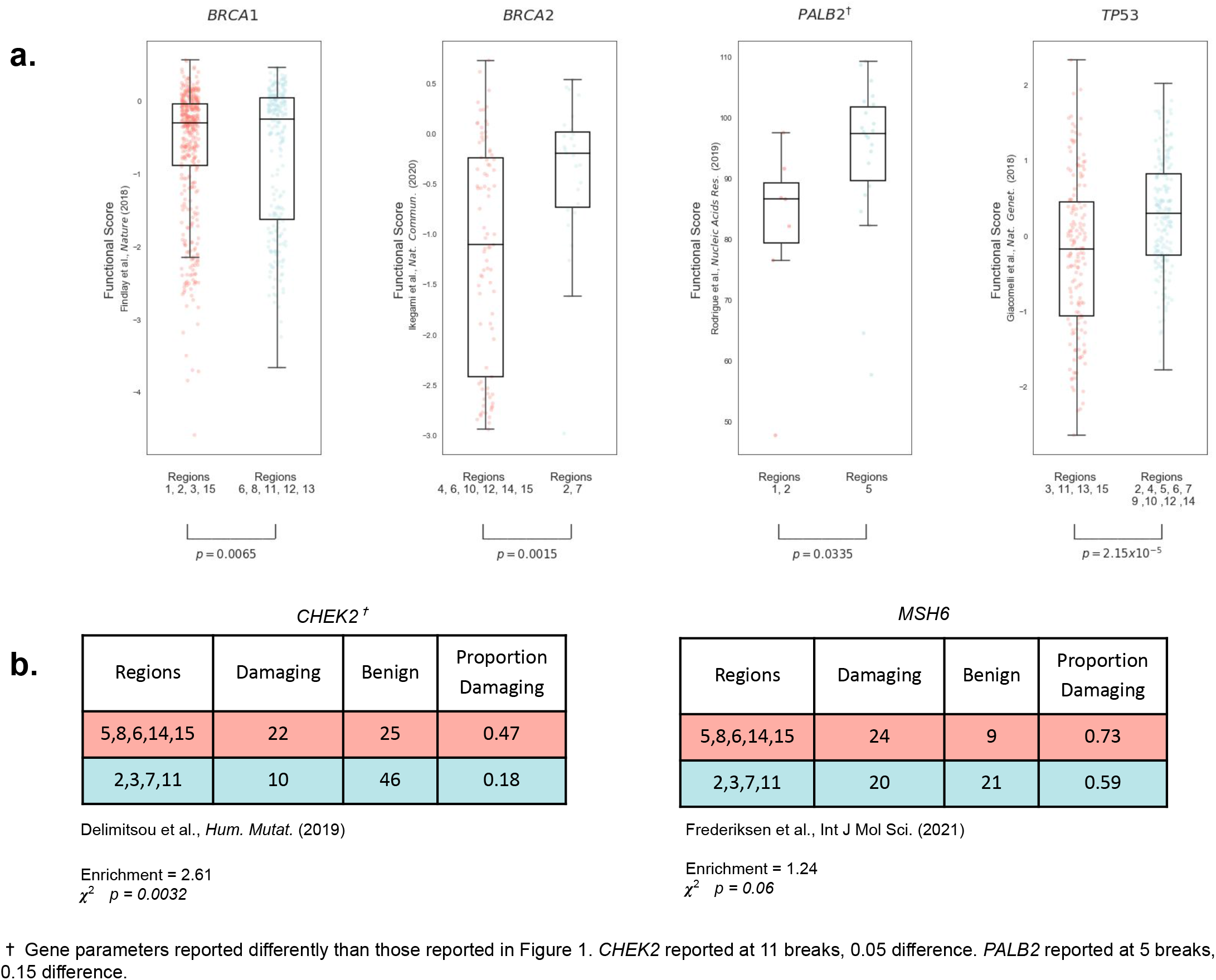
Validation of regional assignments using variant functional assays. **[a]** When comparing functional estimates of variant impact from high-throughput experimental assays with assigned high risk regions (HRRs) and low risk regions (LRRs), we find significant differences between functional assay values in each region type. Scatter plots are shown for datasets where functional values are reported on a continuous scale. **[b]** Tables shown for datasets where functional impact is assessed on a binary Damaging/Benign scale.

### Comparison to protein domains and other methods to differentiate risk in regions

We compare alternative approaches to stratify risk in genic regions, starting with annotated protein domains (Pfam).^32^ In most genes, variants collectively within the protein domains confer no significant difference in breast cancer risk when compared to variants outside of protein domains **(Supplementary Table 5)**. Notably in *MSH6*, variants within any domain confer elevated, but non-significant risk (O.R = 1.36 [1.01, 1.72], logrank p = 0.236). In this case, *MSH6* Pfam domains substantially overlap with REGatta HRRs, comprising 75.5% of the HRR coding positions and 87.8% of HRR carriers **(Supplementary Figure 3)**. In comparison, our regional approach identifies a significant difference between carriers in HRR vs. LRR segments with a larger mean effect size (O.R. = 1.61 [1.23, 1.99], logrank p = 0.008).

Among the 21 domains across these genes, only three confer significantly increased risk, and all three overlap considerably with our defined HRRs. Two of these domains are in *MSH6* (MutS domain I 62.75% overlap, MutS Domain V 54.3% overlap), and the third is the *BRCA1* RING domain. The 40 amino acid RING domain confers the highest O.R. of any domain (O.R. = 5.86 [5.87, 6.86], logrank p = 9.32×10^−7^), and is located entirely in *BRCA1* Regions 1 and 2, the two highest risk regions in *BRCA1*. We find that this elevated clinical risk appears to extend beyond the RING domain: carriers of missense variants in *BRCA1* regions 1 and 2 *outside* of the RING domain have an increased effect size when compared to non-carriers (n=33 individuals, O.R. = 2.51 [1.64, 3.37], logrank p = 0.09). The *ATM* FAT domain is the next most significant domain by logrank p-value (p = 0.056), which has complete overlap with HRRs in *ATM*. HRR carriers outside this domain also have significantly elevated breast cancer incidence vs. non-carriers (O.R. = 1.39, [1.15, 1.63], logrank p = 0.0039) (**Supplementary Figure 3, Supplementary Tables 2, 5**).

Prior work has made use of the clustering of ClinVar reports to inform risk predictions for missense variants.^33^ We compare to one such method which can effectively discriminate between known pathogenic and benign variation using this information (MutScore), and has identified non-random distributions of such variation in regions in a broad set of 559 genes with clinical associations. We apply this score to population cohort data in the 7 genes analyzed, where our method can identify regions with significantly higher and lower risk, and we find no significant difference in breast cancer incidence among those in the top third vs bottom third of MutScore values (**Supplementary Table 6**), the thresholds used in that study.

### Sensitivity analysis for optimization of parameters by gene

Finally, we assess whether the parameter space for numbers of genic regions and effect size thresholds may be optimized per gene, rather than using a fixed value of ± 0.15 for HRR/LRR and a fixed (or maximum) number of regions. Different parameter values yield models with more significant associations in certain genes, as measured by logrank p-value (**Supplementary Figure 4**). We also find that all seven genes have at least one significant result by Benjamini-Hochberg correction (α = 0.01) controlling for combinations of parameters (**Supplementary Table 2**), demonstrating the robustness of this approach to choice of model parameters. We also identify significant differences in variant functional scores under a variety of region and risk threshold parameter configurations (**Supplementary Table 4**).

## Discussion

Germline risk assessment for cancer syndromes is a major application area of precision medicine.^34^ Population sequencing cohorts have sufficiently expanded to identify pathogenic variants associated with clinical outcomes.^35^ However, it is still challenging to estimate the clinical risk rare missense variants generally confer, given limited numbers of observations of each variant and many with smaller effect sizes. We approach this problem by measuring the clinical impact of missense variation within predefined regions from a large national biobank, which provides a uniform assessment of clinical risk.

This provides an attractive alternative to estimating the strength of purifying selection in regions from patterns of variation present in the general population. While methods to assess selective constraint can be a powerful predictor of pathogenicity and can be sensitive in regions under strong selection, they can be limited in resolution for missense variation for genes under weaker selection, due to stochastic effects of drift. Alternative regional boundaries, such as protein domains, may be limited in their applicability due to small size or low numbers of variant carriers, potentially leading to overdispersed estimates of effect, and may also miss putatively damaging variation outside of known protein domains. Finally, methods which provide estimates of functional effects using the relative abundance of pathogenic and/or neutral variation may provide biased estimates of functional effect, as they are not derived from neutrally ascertained populations. For example, the lack of known pathogenic variant reports has been used to argue that variants within certain genic regions are unlikely to be pathogenic, including a large ‘cold spot’ within exon 11 of *BRCA2*.^36^ This conflicts with high risk regions identified by REGatta in *BRCA2*, where regions 4 and 6 fall within a “cold spot” encompassing exon 11, and confer significantly increased risk.

The estimates of effect size that we have produced for carriers of variants in HRRs may serve as a useful prior for population-level risk, which may be useful in diagnostic variant assessment. The ACMG/AMP sequence variant interpretation guidelines consider many sources of evidence in favor of pathogenicity, including computational predictions of variant impact (PP3), presence in a known protein domain (PM1), experimental assays of functional impact (PS3), or absence in population databases (PS4), which may overlap with the evidence defined by REGatta. This approach makes use of newly abundant population data linked with clinical outcomes to infer which regions may be associated with elevated clinical risk.

Limitations of this work include population ascertainment of the UKB, which may bias estimates of effect size, due to demographics and genetic architecture. Though we are currently limited by a small set of genes, these are some of the most commonly screened genes in the diagnostic setting associated with predispositional cancer risk.^37^ Given that we are making estimates from germline variation, it is worth noting that germline sequencing may uncover somatic variants associated with clonal hematopoiesis (CH), a process that occurs more frequently in older individuals. These putatively somatic variants arising from this process have been shown to be pathogenic.^38^ These variants may be filtered by variant allelic fraction, but it may be imperfect to effectively differentiate between somatic and germline variants in older individuals.

Future work includes integrating these regional risk ratios with computational or experimental predictions of functional effect at the variant level, potentially in concert with individual-level risk factors (e.g., family history, lifestyle and behavioral risk factors, and polygenic risk scores).^39^ Additional studies may also assess the actionability of these risk assessments as they may help optimize choice of therapy (e.g., PARP inhibitors) in *BRCA1* or *BRCA2* carriers. Additional work should include expansion to additional genes and phenotypes with strong associations for rare coding variation.

## Data Availability

All data produced in the present study are available upon reasonable request to the authors and with approval from the relevant governing organizations.

https://github.com/cassalab/regatta

## Acknowledgments

We are indebted to the UK Biobank and its participants who provided biological samples and data for this analysis. Work was performed under UK Biobank application #41250 and Mass General Brigham IRB protocol 2020P002093. We gratefully acknowledge helpful advice from Drs. Peter Kraft, Matthew Lebo, Natasha Strande, and Shamil Sunyaev.

## Supplementary Methods

### Study design, setting and participants

The UKB is a prospective cohort of over 500,000 individuals recruited between 2006 and 2010 of ages 40-69.^19^ Drawing from 200,625 participants with exome sequencing data were included in this analysis, we analyze 109,581 female participants. Analysis of the UKB data was approved by the Mass General Brigham Institutional Review Board (Protocol 2020P002093). Work was performed under UKB application #41250.

### Clinical endpoints

The primary clinical endpoint is breast cancer, and case definitions were defined in the UKB using a combination of self-reported data confirmed by healthcare professionals, hospitalization records, and national procedural, cancer, and death registries, previously described at the disorder level.^40^

### Filtering variants used in regional boundary generation

For generating variants used in regional partitioning lists, we limit single nucleotide variants that are not expected to affect splicing, protein length, or that are reported to confer any consequence beyond missense variation. We limit to reports specified as pathogenic or likely pathogenic. We count multiple submissions of the same variant as multiple points in the final list to generate the regional boundaries.

### Statistical tests and software

Statistical tests are two-sided unless otherwise reported. Statistical tests were performed using scipy v1.4.1 for KS and ^2^ tests.^41^ Logrank tests, KM curve figures, and Cox proportional hazard regressions were performed using lifelines v0.25.2.^42^ Additional analyses were performed using Numpy v1.17.3, Pandas v.1.1.5, and scikit-learn v.0.24.2.^43–45^ Protein structure images created with PyMol v2.5.2.^46^ Additional images were created using matplotlib v3.1.1 and seaborn v0.9.0.^47,48^

### Functional Analyses

In the included high throughput variant installation assays, some studies reported cellular phenotypic values on a continuous scale while others reported these results in some form of binary classification. When multiple amino acid changes are reported for the same reference amino acid we use the average of all available measurements to represent each position. We then take the average of each regions’ positional scores to represent the regional value. No processing on the data was done outside of that originally reported by the authors’ data.

### Mutscore comparison

We limited our analyses to missense variants reported in the dataset. We defined high risk variants as those reported with a score ≥ 0.66 and low risk variants as those with a score ≤ 0.33. We select variant carriers in the same fashion we did for the remaining analyses. Data was obtained from the online repository https://mutscore-wgt7hvakhq-ew.a.run.app/.

### Data and code availability

Code used for all analyses and all figure creation is available at https://github.com/cassalab/regatta. The publically available data underlying these analyses (Pfam domains, ClinVar, MutScore, functional data, and AlphaFold Structural Predictions) are available in annotated files in the repository as well.

---

**Supplementary Figure 1.**
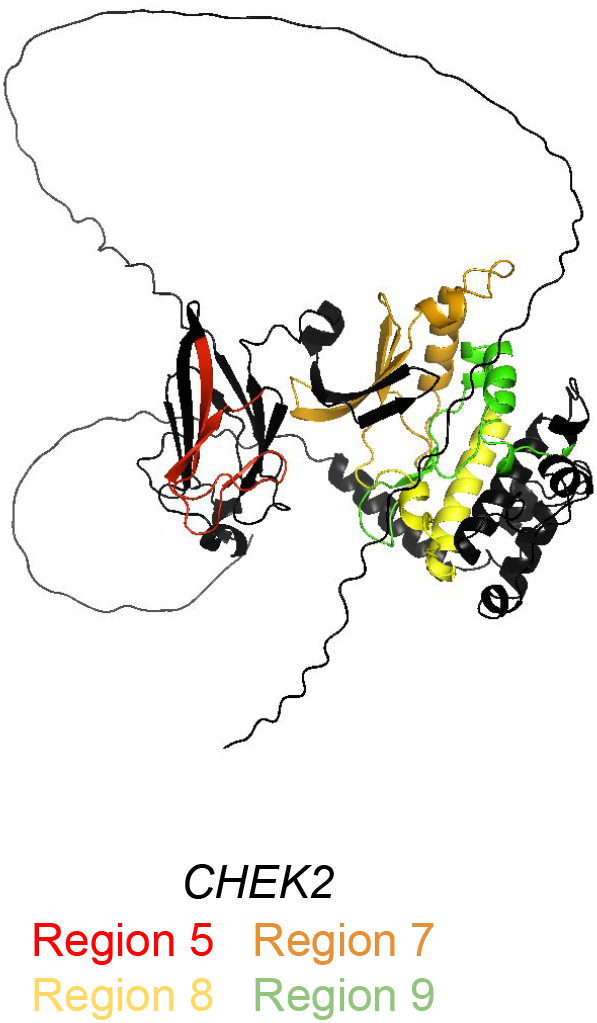
Spatial assessment of HRRs. AlphaFold predicted protein structures with predicted high risk regions, including *CHEK2* which was not included in Figure 2.

**Supplementary Figure 2.**
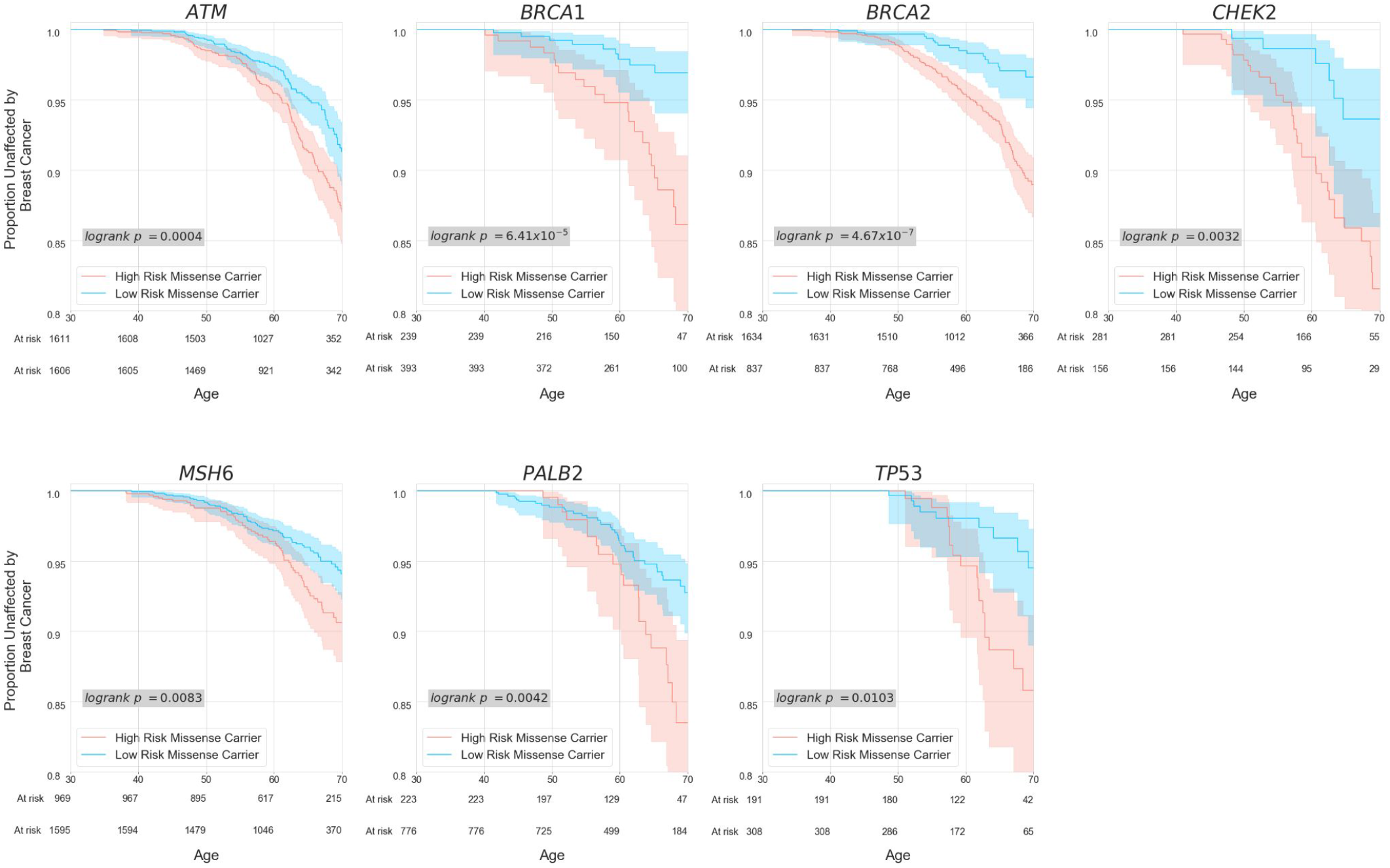
Kaplan-Meier curves comparing risk for carriers of missense variants in HRRs vs. LRRs. Regional assignments are defined in **Figure 2**.

**Supplementary Figure 3.**
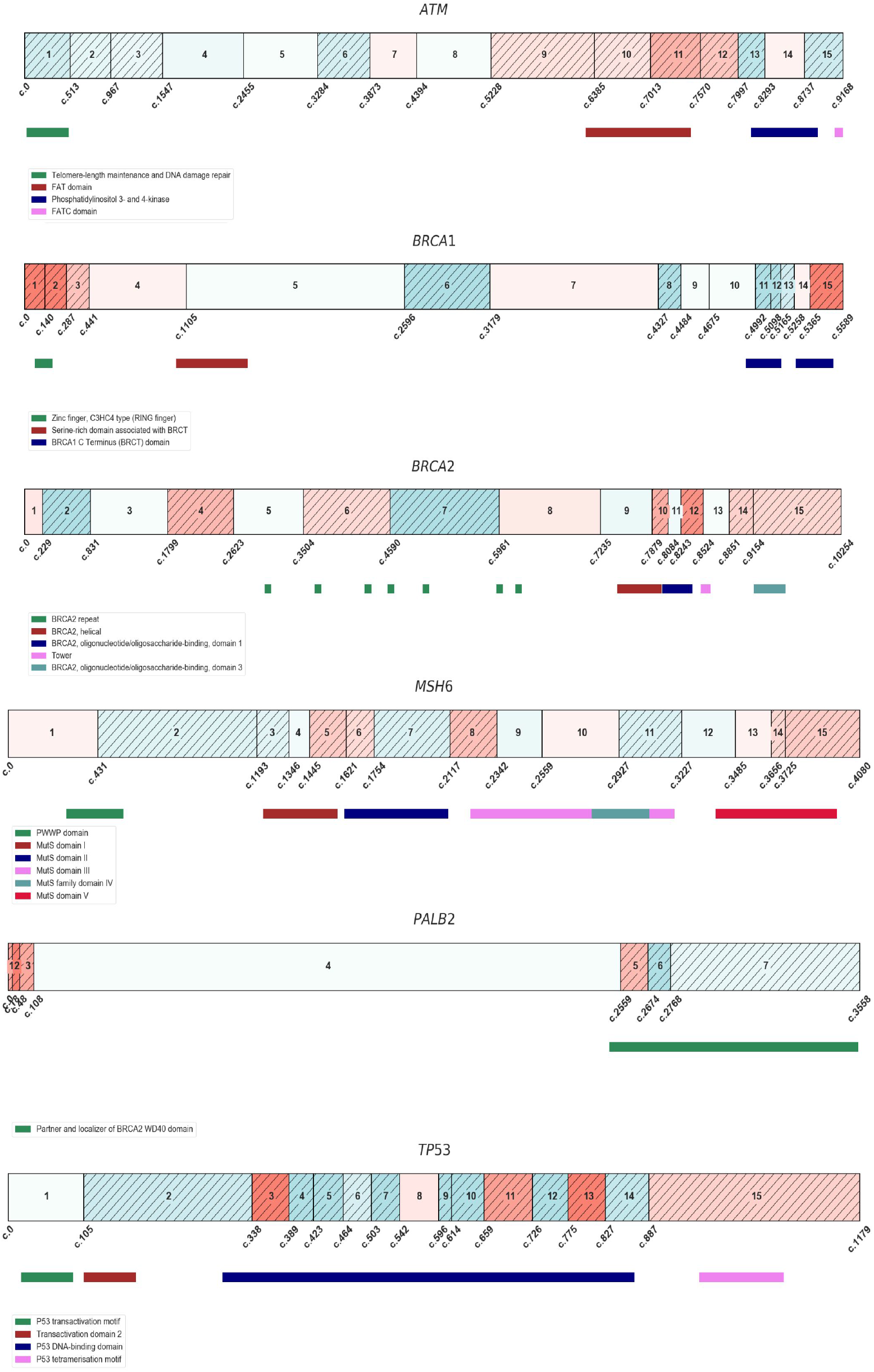
Pfam domains among regions of high and low risk. Among the genes analyzed, *MSH6* was the only gene where missense variants within annotated Pfam domains conferred additional risk over missense variants outside of Pfam domains. Here, Pfam domains overlap considerably with the two *MSH6* HRRs (domains cover 56% of HRR positions in the gene). Individual O.R. for each domain can be found in Supplementary Table 5.

**Supplementary Figure 4.**
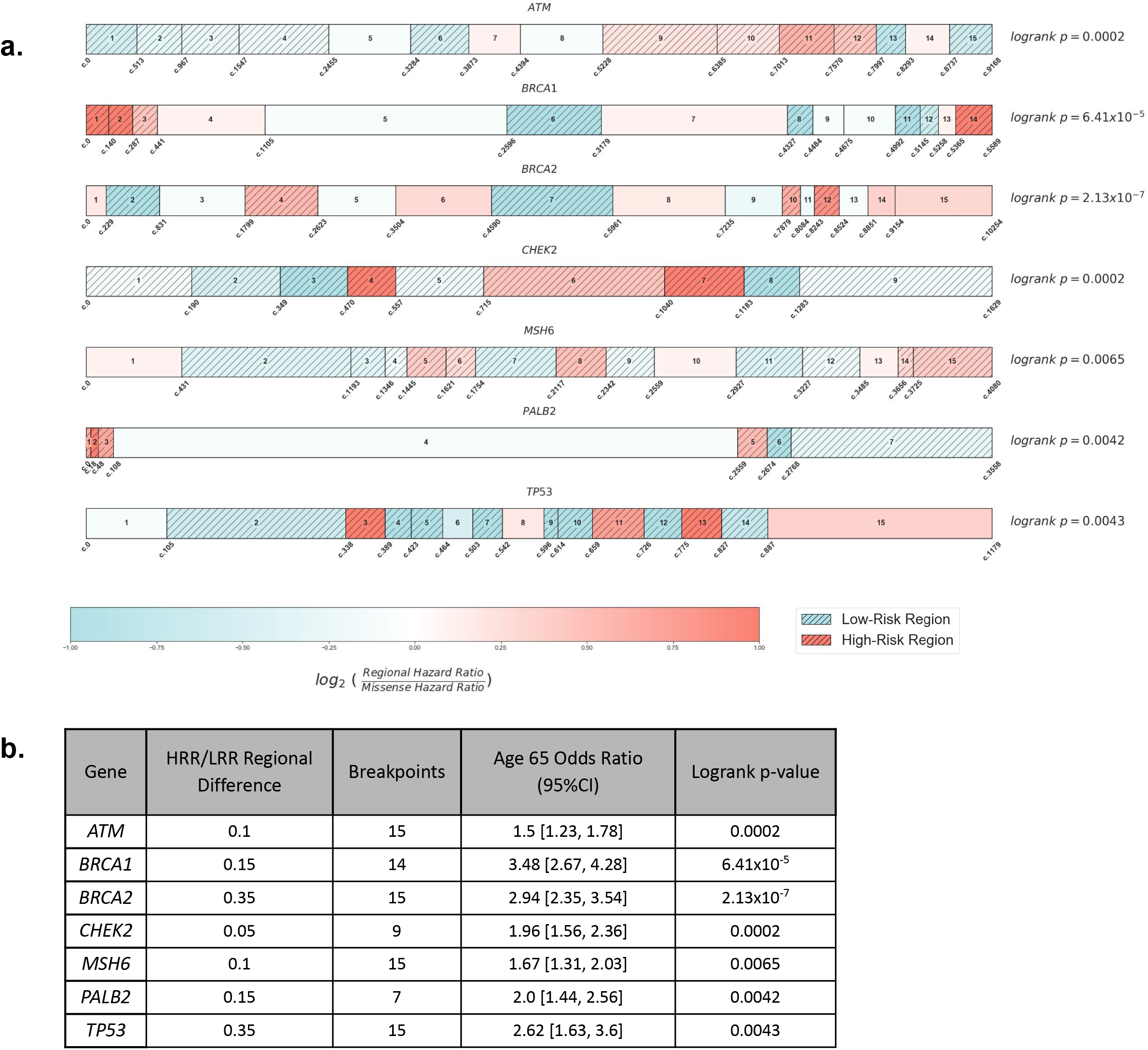
Maximizing differences in breast cancer outcomes through regional partitions. **a)** Diagram of differential risk by gene when selecting the optimal configuration of regional difference and number of breaks. Optimal configuration is determined based on the minimum logrank p-value. **b)** Further information regarding the optimum configurations of parameters, and their associated breast cancer outcome data.

## Explanation of Supplementary Tables

Supplementary Table 1 - Regional regression results from each individual region tested when making regional assignments. Output of the Cox proportional hazard model is shown for each region in each configuration of parameters tested.

Supplementary Table 2 : Logrank p-values and regional assignments from all configurations of parameters test.

Supplementary Table 3 - Data underlying the odds ratios shown in Figure 3A for all groups considered in the odds ratio comparisons.

Supplementary Table 4 : Functional results of all tested configurations of partitions.

Supplementary Table 5 : Comparisons were done between non-carriers of coding variants and individuals with missense variants in each domain. Domains where RR or logrank p-value calculation could not be done are not shown. This is due to lack of carriers and/or breast cancer cases among carriers in those domains. Coverage calculations come from breakpoint configurations reported in Figure 2A.

Supplementary Table 6 : Data comparing BCAC study genes breast cancer outcomes in utilizing a previously described method which also relies on ClinVar reports of pathogenicity. (Quinodoz, M. et al. The American Journal of Human Genetics (2022))

